# A Deep Learning Pipeline for Analysis of the 3D Morphology of the Cerebral Small Perforating Arteries from Time-of-Flight 7 Tesla MRI

**DOI:** 10.1101/2024.10.03.24314845

**Authors:** Rui Li, Soumick Chatterjee, Yeerfan Jiaerken, Chethan Radhakrishna, Philip Benjamin, Stefania Nannoni, Daniel J. Tozer, Hugh S. Markus, Christopher T. Rodgers

**Affiliations:** Department of Clinical Neurosciences, University of Cambridge, Cambridge, United Kingdom; Genomics Research Centre, Human Technopole, Milan, Italy; Faculty of Computer Science, Otto von Guericke University Magdeburg, Magdeburg, Germany; Department of Radiology, Second Affiliated Hospital of Zhejiang University School of Medicine, Zhejiang, China; Atkinson Morley Regional Neuroscience Centre, St George’s University Hospitals NHS Foundation Trust, London, United Kingdom; Wolfson Brain Imaging Centre, Department of Clinical Neurosciences, University of Cambridge, Cambridge, United Kingdom

## Abstract

The lenticulostriate arteries (LSA) supply important subcortical structures in the brain and are affected in cerebral small vessel disease (CSVD), leading to changes in their morphology. 7 Tesla Time-of-Flight magnetic resonance angiography (7T-TOF-MRA) now allows their visualisation in humans, but current analysis of LSA morphology largely relies on manual tracing on 2D coronal maximum-intensity-projection (MIP) images, which discards significant information from the third spatial dimension. We aimed to develop a semi-automatic pipeline for quantifying the 3D morphology of LSAs from 7T-TOF-MRA in patients with CSVD.

We used contrast-enhanced 7T-TOF-MRA data from 15 subjects enrolled in a local CSVD study. Our pipeline consists of two main stages: vessel segmentation and LSA quantification. For segmentation, we fine-tuned a state-of-the-art deep learning model, “DS6”, for vessel segmentation and compared its performance against a classical Frangi filter-based pipeline, Multi-Scale Frangi Diffusive Filter (MSFDF). Both methods were evaluated against manually labelled ground-truth masks in LSA regions. In the LSA quantification stage, the user defines a region-of-interest around LSAs and checks the segmentation. Based on this, the LSA centrelines are extracted, and branch counts, length, tortuosity, and curvature are computed. Additionally, we conducted the traditional LSA analysis using 2D coronal MIPs, and we evaluated the correlation between the results from the 2D and 3D analyses.

For vessel segmentation, the fine-tuned DS6 model achieved a mean Dice similarity coefficient (DSC) of 0.814±0.029 during testing, outperforming MSFDF on DSC, sensitivity, and balanced average Hausdorff distance in terms of both mean value and stability. Visual inspection confirmed that DS6 was more sensitive in detecting LSA branches with weak signals. On average, the 15 subjects had 5.9±1.6 LSA stems and 28.7±9.9 branches. The mean length of an LSA branch was 42.5±5.7mm, and mean tortuosity was 1.9±0.2. Finally, the branch counts from 2D and 3D analyses correlated well (*ρ*=0.741, *p*=2.816e-06), whereas the stem count, branch length and tortuosity measurements were significantly different, showing the insufficiency of MIP analysis (stem: *ρ*=0.230, *p*=2.207e-01; length: *r*=0.565, *p*=1.153e-03; tortuosity: *r*=0.400, *p*=2.847e-02).

We have developed an open-source semi-automatic pipeline using deep learning for evaluating the 3D morphology of LSAs in CSVD patients from 7T-TOF-MRA. We show that analysing LSA morphology in 3D reveals previously inaccessible aspects of morphology. Our pipeline offers a valuable tool for clinical research studies to characterise the 3D morphology of LSAs in CSVD.

## 1. Introduction

The lenticulostriate arteries (LSA) are a group of small perforating arteries in the brain that originate from the proximal segments of the middle cerebral artery (MCA) and anterior cerebral artery (ACA). They supply blood to important subcortical structures, including the basal ganglia and internal capsule [1, 2]. The LSAs are implicated in the pathogenesis of cerebral small vessel disease (CSVD) – a cerebrovascular disorder causing 25% of ischaemic strokes, most haemorrhagic strokes, and it is the major pathology underlying vascular dementia [3, 4]. CSVD can cause the narrowing, occlusion, or rupture of LSAs, which may eventually lead to lacunar stroke or intracerebral haemorrhage [5, 6]. Therefore, evaluating changes to the LSAs in CSVD patients may detect early pathological processes and offer a valuable perspective on the disease mechanisms of CSVD.

Due to the small size of LSAs (0.10-1.28 mm in diameter) [7], in vivo visualisation of LSAs has been challenging until the advent of 7 Tesla time-of-flight magnetic resonance angiography (7T-TOF-MRA), which provides higher spatial resolutions than clinical 1.5/3T MRI [8]. Using a gadolinium contrast agent further enhances the visibility of LSAs [9]. These advances in imaging have prompted the emergence of many clinical studies examining LSAs using 7T MRA [9–13]. However, these studies produce hundreds of high-resolution slices which are burdensome to assess manually in 3D. There is a clear need for a reliable (semi-)automated tool to make it practical to evaluate the morphology of LSAs in 3D in clinically meaningful numbers of patients. We aimed to develop the needed pipeline for analysis of the 3D morphology of LSAs from 7T-TOF-MRA in patients with CSVD.

### 1.1. Background and Related Work

In the current clinical literature, the most popular method for evaluating LSA morphology is using coronal maximum intensity projection (MIP) images from TOF data covering the LSA region (or minimum intensity projection images for “dark-blood” imaging such as vessel-wall imaging) [2, 9, 10, 14–17]. As illustrated in Supplementary Materials Figure 1, on this 2D projection, the clinician manually delineates the longest LSA branch or all LSA branches on each side of the brain, from which metrics like apparent length and tortuosity are reported. This method is relatively simple to use, but the major disadvantage is that the 2D projection cannot fully reflect the 3D structure of LSAs. Moreover, branches can “cross” on the MIP (while not in 3D), and the bottom parts of the LSAs are often occluded by the MCA/ACA. Together, these factors make LSA morphology extracted from MIPs less reliable.

Therefore, it is desirable to extract the morphology of LSAs in 3D, and some studies have attempted this. Liao *et al.* [18] developed a model-based segmentation approach, in which, after initialising a starting point on a vessel centreline, their algorithm incrementally fits 3D parametric tube models based on voxel intensities in the TOF volume. The fitted tubes thus trace the vessels, and the centrelines and radii of the vessels are automatically extracted from the fitted parameters. A minimal path algorithm is used to fill any gaps in the initial segmentation in areas with low signal or noise/artefact. They applied this approach to segment LSAs from 7T-TOF-MRA in patients with vascular dementia [13]. However, their study did not evaluate segmentation accuracy quantitatively. Their method is also not publicly available, which makes it unsuitable for our use case.

Most other methods for analysing vessel morphology in 3D require a two-stage approach: first vessel segmentation, then morphology quantification with centreline extraction, bifurcation detection, and construction of the vessel tree. Many vessel segmentation methods have been proposed [19], and one of the most classical and popular approaches is multiscale vessel enhancement filtering using Frangi filters [20]. It applies a Hessian filter to the raw images to enhance tubular structures at multiple diameters while suppressing background noise. The filtered images are then often thresholded to achieve binary segmentation.

Bernier *et al.* [21] applied an extension of the Frangi filter to segment human cerebral vasculature from both 3T TOF MRA and susceptibility weighted images (SWI) in a cohort of young healthy volunteers. Their open-source analysis pipeline, named Multi-Scale Frangi Diffusive Filter (MSFDF), has also seen many applications afterwards [22–24]. Similarly, Liu *et al.* [25] combined modified Frangi filtering with automatic seed detection and region growing techniques to reduce the false positive errors in segmentation. They developed a pipeline for the comparative analysis of the cerebral vasculature in young healthy volunteers using both 3T and 7T TOF MRA data. Moreover, Deshpande *et al.* [26] utilised Frangi filter-based segmentation for the comparative analysis of the vasculature between healthy volunteers and stroke patients, using both 1.5T TOF MRA and computed tomography angiograms (CTA). In particular for LSAs, Wei *et al.* [11] applied vessel enhancement filtering to segmenting LSAs and quantified their volume inside the basal ganglia. However, no evaluation of the segmentation accuracy was reported, and they resorted to using manual labelling on 2D coronal MIPs for measuring LSA apparent length.

In recent years, deep learning (DL) methods have seen increasing success in vessel segmentation. Livne *et al.* [27] conducted the first study utilising the 2D U-Net architecture [28] to segment cerebral arteries from 3T TOF MRA in patients with cerebrovascular disease. They performed segmentation on 2D patches cropped from slices of the 3D volume, achieving high accuracy compared to manually labelled ground truth. To tackle the difficulty in manually annotating a large amount of training data, Fan *et al.* [29] proposed an unsupervised scheme where the TOF volumes are initially segmented by a hidden Markov random field model, and the masks are subsequently used as ground truth to train another deep learning segmentation model like U-Net. Furthermore, Chatterjee *et al.* [30] proposed the “DS6” model, incorporating a multi-level deep supervision scheme for 3D U-Net [31] with elastic deformation consistency learning [32]. Their approach requires only a small amount of training data (6 MRA volumes, cropped into 20,412 patches) and outperformed the original U-Net, attention U-Net and the MSFDF pipeline. Outside U-Net, Tetteh et al. [33] proposed a fully convolutional network for vessel segmentation using 3D cross-hair filters to enhance memory usage and speed.

A summary of these papers is provided in Supplementary Materials Table 1. However, most of these papers on vessel segmentation did not specifically focus on small vessels but rather the whole-brain vasculature [21, 25–27, 29, 30, 33]. Their evaluations were therefore dominated by large vessels due to their large volume contributing more pixels and hence dominating DSC metrics. The performance of these models remains to be tested on small vessels like LSAs, which are by definition narrower, and also have much weaker and less homogeneous signals than large vessels. Additionally, many models were developed using MRA data from young healthy volunteers [21, 25, 30], which typically provide higher quality images with better signal-to-noise than those acquired from elderly patients with CSVD. Thus, these models’ segmentation performance on CSVD patient data remains unclear. Furthermore, few studies have utilised contrast enhanced TOF, which can better visualise LSAs [9].

**Table 1.** Demographical and clinical data of selected subjects. Age and BMI are shown as mean ± standard deviation. BMI=Body Mass Index.

|  |  |
| --- | --- |
| **N** | 15 |
| **Group** | Symptomatic CSVD: 11 (73.3%)<br>Healthy Control: 2 (13.3%)<br>Non-lacunar Stroke: 2 (13.3%) |
| **Age in years** | 62.5 ± 11.9 |
| **Sex** | Male: 8 (53.3%)<br>Female: 7 (46.7%) |
| **Ethnicity** | Asian or Asian British: 1 (6.7%)<br>White: 13 (86.7%)<br>Unknown: 1 (6.7%) |
| **Body mass index** | 28.8 ± 4.4 |
| **Smoking** | Never Smoked: 5 (33.3%)<br>Ex-Smoker: 7 (46.7%)<br>Current Smoker: 3 (20.0%) |
| **Drink Alcohol** | 9 (60.0%) |
| **Hypertension** | 8 (53.3%) |
| **Hyperlipidaemia** | 6 (40.0%) |
| **Diabetes Mellitus** | 2 (13.3%) |
| **Ischemic Heart Disease** | 1 (6.7%) |
| **Atrial Fibrillation** | 2 (13.3%) |
| **Previous Stroke** | 2 (13.3%) |

Furthermore, only a few papers developed methods for vessel morphology quantification after segmentation [21, 25, 26], and most obtained the centreline skeleton using morphological thinning, which is sensitive to unsmooth segmentation boundaries and tends to produce spurs and loops on the centrelines [34]. This can be particularly problematic for the analysis of LSAs, where we are interested in the tortuosity of the vessel centrelines. Thus, more robust centreline extraction algorithms are desired.

### 1.2. Contribution

This paper proposes a deep learning-based pipeline for the analysis of the 3D morphology of LSAs in CSVD patients from contrast enhanced 7T-TOF-MRA data. Our pipeline consists of an automatic stage of vessel segmentation, and a semi-automatic stage to extract vessel centrelines and compute morphological metrics. For vessel segmentation, we applied a state-of-the-art deep learning model, DS6, having finetuned it to enhanced its performance on data from CSVD patients. To our knowledge, this is the first application of deep learning models to the segmentation of LSAs, and we also compared its performance with the popular classical approach, the MSFDF pipeline. For LSA morphology quantification, we developed an interactive pipeline integrating functionalities from both Python and the 3D Slicer software, which we present as a Python Jupyter Notebook for ease of use. Our pipeline is open source to facilitate its application in clinical research on cerebrovascular disease.

## 2. Materials and Methods

### 2.1. Data

We used data from the Cambridge 7T Cerebral Small Vessel Disease (CamSVD) study [9], which is an ongoing 7T MRI study recruiting participants from the following 3 categories:

i. Symptomatic CSVD: defined as a lacunar stroke syndrome with an anatomically corresponding lacunar infarct on MRI.
ii. Healthy volunteer controls: age matched healthy volunteers with no evidence of major neurological diseases.
iii. Non-lacunar strokes without CSVD: patients with stroke due to occlusion of the larger cerebral arteries and not caused by CSVD.

Detailed inclusion and exclusion criteria for each group are provided in Supplementary Materials Table 2. All stroke patients were past the acute phase. Written informed consent was obtained from all participants. The CamSVD study was approved by the Institutional Review Board of East of England–Cambridge Central Research Ethics Committee (REC Ref: 19/EE/0219). For the development and evaluation of our pipeline, we randomly selected 15 subjects with satisfactory image quality (e.g., no severe head motion artefact, so that LSAs are visible) from the current cohort of CamSVD.

**Table 2.** Quantitative evaluation results of the original and fine-tuned DS6 model and the tuned MSFDF pipeline. For metrics with ↑, the higher the value the better. For metrics with ↓, the lower the value the better. For fine-tuned DS6, values in the *mean Δ* columns show the mean percentage difference compared with the original DS6. For MSFDF, values in the *Mean Δ* columns show the mean percentage difference compared with the fine-tuned DS6. DSC=Dice similarity coefficient; bAHD=balanced Average Hausdorff Distance; PPV=Positive Predictive Value.

| Model | Status | Dataset | DSC ↑ | | bAHD ↓ | | Sensitivity ↑ | | PPV ↑ | |
| --- | --- | --- | --- | --- | --- | --- | --- | --- | --- | --- |
|  |  |  | *Mean ± SD* | *Mean $\Delta$* | *Mean ± SD* | *Mean $\Delta$* | *Mean ± SD* | *Mean $\Delta$* | *Mean ± SD* | *Mean $\Delta$* |
| DS6 | Original | Training | 0.811 ± 0.057 | | 0.480 ± 0.246 | | 0.867 ± 0.088 | | 0.768 ± 0.070 | |
| | | Validation | 0.780 ± 0.053 | | 0.605 ± 0.184 | | 0.817 ± 0.071 | | 0.746 ± 0.041 | |
| | | Test | 0.767 ± 0.031 | | 0.619 ± 0.205 | | 0.811 ± 0.079 | | 0.733 ± 0.036 | |
| DS6 | Fine-tuned | Training | 0.846 ± 0.035 | + 4.69% | 0.316 ± 0.139 | − 30.26% | 0.892 ± 0.064 | + 3.36% | 0.807 ± 0.034 | + 5.67% |
| | | Validation | 0.800 ± 0.030 | + 2.92% | 0.490 ± 0.099 | − 15.38% | 0.860 ± 0.043 | + 5.90% | 0.751 ± 0.042 | + 0.58% |
| | | Test | 0.814 ± 0.029 | + 6.27% | 0.421 ± 0.142 | − 30.94% | 0.863 ± 0.045 | + 6.96% | 0.772 ± 0.029 | + 5.55% |
| MSFDF | Tuned | Tuning | 0.777 ± 0.065 | − 6.58% | 0.710 ± 0.733 | + 75.91% | 0.748 ± 0.065 | − 15.11% | 0.819 ± 0.101 | + 3.73% |
| | | Test | 0.769 ± 0.053 | − 5.53% | 0.576 ± 0.229 | + 50.15% | 0.699 ± 0.083 | − 19.05% | 0.865 ± 0.036 | + 12.00% |

#### 2.1.1. Subject Demographics

The demographics and clinical information of the 15 subjects included are summarised in Table 1. Most 11(73.3%) of the subjects were symptomatic CSVD patients, while 2 each were from the healthy control and non-CSVD strokes group. The mean age was 62.5±11.9, and 53.3% of the subjects were male. Most subjects were white by ethnicity. Mean body mass index (BMI) was 28.8±4.4, which is in the “overweight” category (BMI 25-29.9). 66.7% of the subjects had smoking history, and 60% were alcohol-drinker. Among the other cardiovascular risk factors, hypertension and hyperlipidaemia were present in about 50% of the subjects, while other factors were less prevalent.

#### 2.1.2. MRI Acquisition

Images were acquired on a 7T MAGNETOM Terra MRI (Siemens, Erlangen, Germany). Details of the imaging protocol have been published previously [9]. Of interest for this study was the 7T-TOF-MRA sequence. A 20 gauge intravenous cannula was inserted in the antecubital vein, and 0.1 mmol/kg of a gadolinium-based contrast agent (Gadobutrol, Gadovist®, Bayer PLC, Reading, UK), followed by 10 millilitres of 0.9% sodium chloride were then administered manually. After a delay of two minutes TOF-MRA data was acquired with the following parameters: field of view (FOV) 200 × 156.3 mm^2^, voxel Size 0.24 × 0.24 × 0.32 mm^3^, repetition time (TR) 13 ms, echo time (TE) 5.1 ms, and nominal flip angle (FA) 20 degrees. Two slabs were used with 80 slices per slab to shorten the time of flight of inflowing blood to increase the signal from this blood. GeneRalized Autocalibrating Partially Parallel Acquisition (GRAPPA) with an acceleration factor of two was used to reduce the scan time to 9 minutes 53 seconds.

### 2.2. Our Pipeline

An overview of our pipeline is depicted in Figure 1. It consists of two main stages: first, a *segmentation* stage, which performs automatic segmentation of the whole vasculature on the complete 3D TOF MRA volume; second, a *quantification* stage, which is a semi-automatic process that extracts the centrelines of the LSAs and compute morphological metrics of interest.

**Figure 1.**
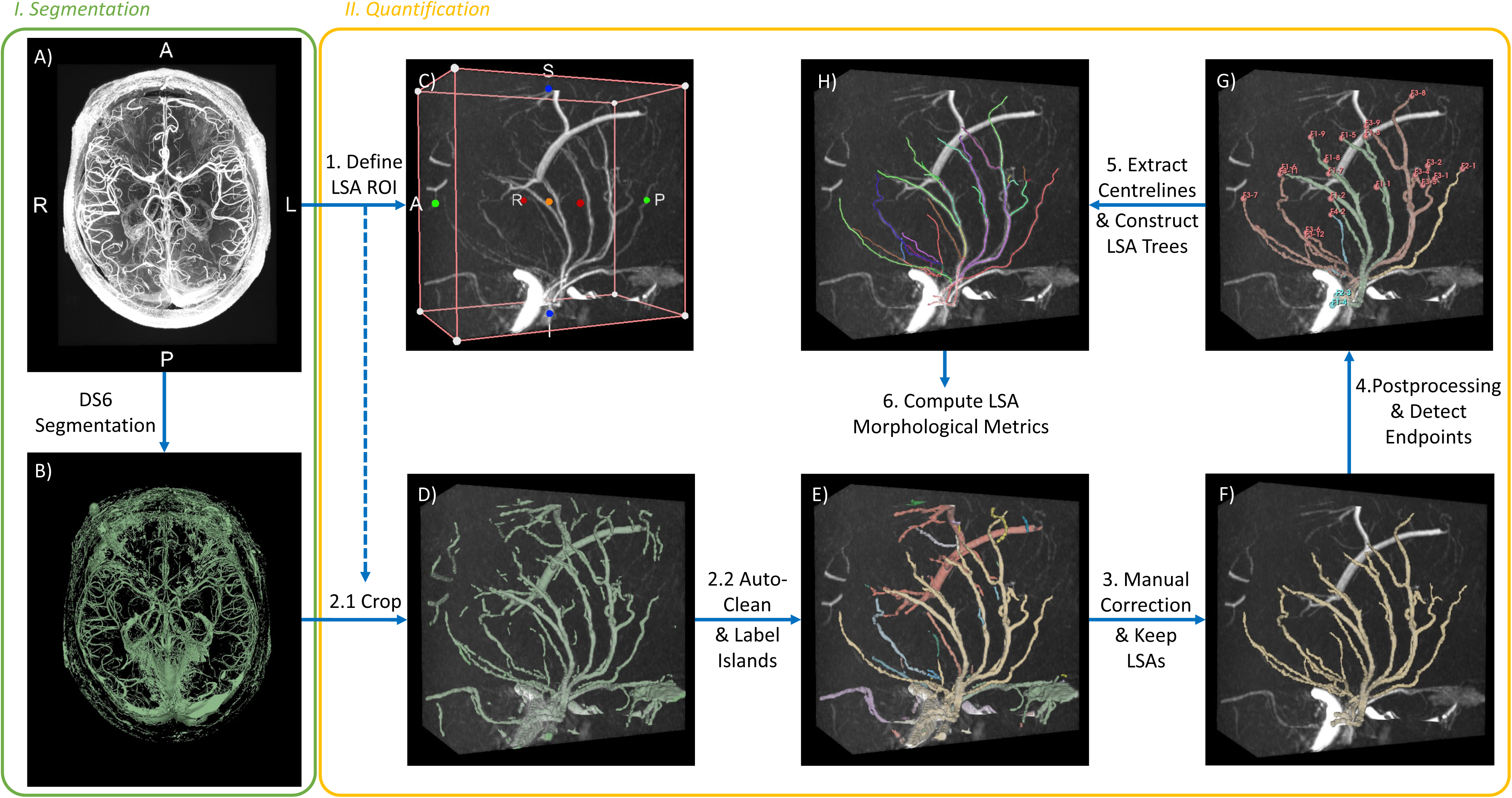
Diagram of the overall proposed pipeline. A) complete TOF volume. B) complete segmentation of the vasculature. C) cropped TOF inside the selected LSA ROI on the left side of the brain. D) cropped segmentation inside the selected LSA ROI, imposed on TOF. E) segmentation after automatic cleaning, with disjoint islands labelled with different colours to assist manual correction. F) manually corrected segmentation of all LSAs. G) segmentation of LSAs originating from different stems and labelled with different colours, along with the detected endpoints. H) extracted centrelines of the LSAs, with different segments labelled in different colours.

#### 2.2.1. Segmentation Stage

Here we applied a state-of-the-art deep learning model for vessel segmentation called “DS6” [30]. For comparison, we also evaluated a non-deep learning method, the Multi-Scale Frangi Diffusive Filter (MSFDF) Pipeline [21], and compared its performance against DS6.

##### DS6

DS6 is a semi-supervised deep learning-based method developed specifically for the segmentation of vasculature from TOF MRA data [30]. It is based on a modified U-Net Multi-Scale Supervision structure, and it utilises a deformation-aware learning strategy during training, which makes the model robust to elastic deformation of the input volumes, thus improving the generalisation of the model even when trained on a small dataset [30]. Segmentation by DS6 was performed on 3D patches of 64×64×64 voxels, and the final segmentation of a TOF volume was constructed by averaging the segmentation on multiple overlapping 3D patches covering the whole volume.

The DS6 model we applied was pre-trained and validated on 14 whole-brain non-contrast-enhanced 7T TOF MRA volumes from healthy volunteers in the StudyForrest dataset [35], using the annotations provided by the SMILE-UHURA challenge [36]. Given the differences in the use of contrast agent, imaging protocol, and population between StudyForrest and our study, we fine-tuned this DS6 model to improve its performance on our data and particularly in the LSA regions.

##### MSFDF

We chose the MSFDF pipeline for comparison because it represents a major class of vessel segmentation methods based on multi-scale vessel-enhancement filtering, and because it is available as open-source and has been applied in several studies [30, 37, 38].

Details of this pipeline can be found in the original publication [21], and we provide a summary diagram of this pipeline in Supplementary Materials Figure 2. Briefly, it filters the TOF MRA volumes with a set of 3D Frangi vesselness filters [20], each of which enhances tubular structure at a specified scale. The set of filter scales span from the minimum to the maximum diameter of the vessels of interest. Thus, vessels of different sizes are highlighted in different filtered images, and by retaining the maximum value at each voxel across all filtered images, we get a final filtered image with all vessels highlighted. Finally, this vessel-enhanced image is binarised through thresholding to obtain a vessel mask.

We adopted the latest code of the MSFDF pipeline at https://github.com/braincharter/vasculature_notebook ^1^. Additionally, we optimised this pipeline by:

- Replacing the original thresholding method based on Otsu’s method [39], which was found to produce “wall artefacts” in the segmentation mask (Supplementary Materials Figure 3), with adaptive Gaussian thresholding [40]. As shown in Equation (1), the latter method computes a threshold at a location (*x*, *y*, *z*) by computing a Gaussian-weighted mean of the image intensity (*I*) in a local neighbourhood (*N*) spanning *D* voxels in each direction. *G*_(*x*,*y*,*z*)_(*i*, *j*, *k*) is a Gaussian function located at (*x*, *y*, *z*) with variance *σ*^2^, *W* = ∑_(*i*,*j*,*k*)∈*N*(*x*,*y*,*z*)_ *G*(*i*, *j*, *k*) is a normalisation term, and *C* is a scalar offset. As the threshold is computed locally, this method is robust to background gradient changes.

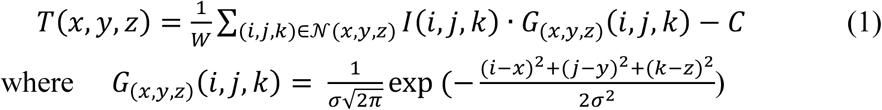

- Postprocessing the segmentation mask by performing morphological closing (the binary morphological structure is a 3×3×3 array with face connectivity) [41], which bridges small gaps, removing small islands distant from other islands, and filling closed holes. This was found to improve its segmentation performance (Supplementary Materials Table 3).

##### Region-of-Interest

As the ultimate goal of our pipeline is to analyse the 3D morphology of LSAs, we were only interested in the models’ segmentation performance on the LSAs. Therefore, for each of the 15 selected subjects from CamSVD, we manually defined a cuboidal region-of-interest (ROI) in each hemisphere containing the cluster of LSAs. Figure 2 illustrates the defined LSA ROIs for an example subject.

**Figure 2.**
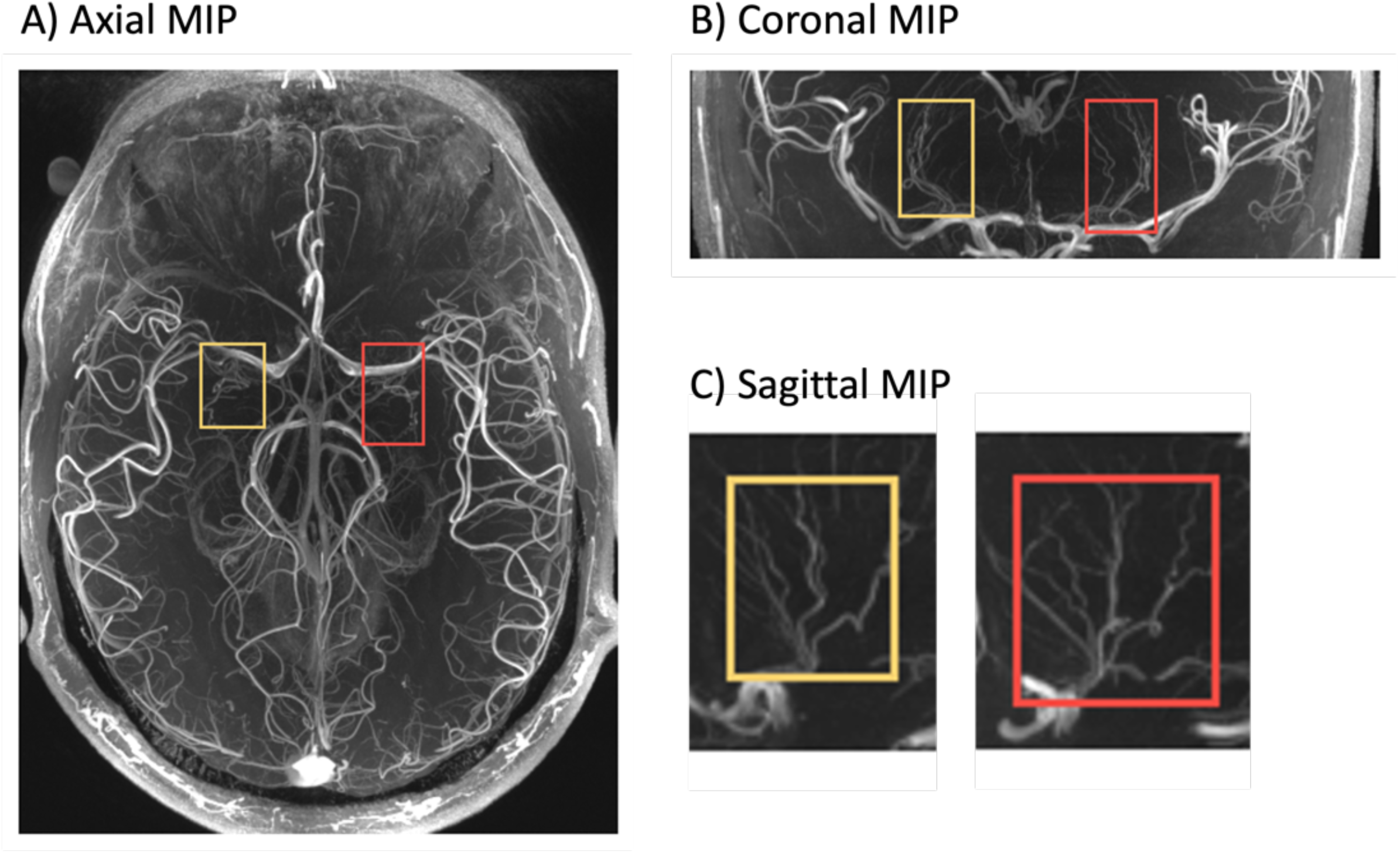
Illustration of the LSA ROIs for an example subject. Left and right (in radiological convention) ROIs are labelled in red and yellow respectively.

**Table 3.** Summary of the extracted LSA morphology metrics from all 15 subjects. Each metric was firstly computed for each subject by averaging over all LSA branches or segments. Reported are the group means and standard deviations (SD) of each subject’s results.

| **Metric** | **Group Mean ± SD** |
| --- | --- |
| Number of Stems | 5.9 ± 1.6 |
| ***Branch*** |  |
| Total number | 28.7 ± 9.9 |
| Mean length (mm) | 42.54 ± 5.68 |
| Mean tortuosity | 1.86 ± 0.15 |
| Mean of average curvature (mm<sup>-1</sup>) | 1.66 ± 0.18 |
| Mean of maximum curvature (mm<sup>-1</sup>) | 7.14 ± 0.54 |
| ***Segment*** |  |
| Total number | 51.3 ± 19.0 |
| Mean length (mm) | 12.74 ± 2.53 |
| Mean tortuosity | 1.32 ± 0.08 |
| Mean of average curvature (mm<sup>-1</sup>) | 1.72 ± 0.18 |
| Mean of maximum curvature (mm<sup>-1</sup>) | 5.75 ± 0.54 |

In principle, when defining the ROI, we aimed to select the smallest box that includes the origins (where the LSAs leave the MCA or ACA) and all branches of the LSAs. However, for some individuals, this was found to include too much of other non-LSA vessel structures (e.g., large vessels, choroid plexus), which could bias the evaluation of the segmentation accuracy for LSAs within the ROI. Therefore, the final LSA ROIs were drawn as a trade-off between the completeness of the LSAs and the non-LSA vessel structures included, and the size of the ROI could be different for different subjects and hemispheres, but they all measured ∼70×100×100 voxels. These LSA ROIs were manually labelled by one trained rater (RL) to obtain the ground-truth masks, which were used for training and evaluating the segmentation models.

##### MRI Preprocessing

Minimal preprocessing was required for the TOF volumes before vessel segmentation. To address the distortion caused by the anisotropic voxel size of the original TOF data (0.24×0.24×0.32 mm^3^), all TOF images were upsampled in the z direction with cubic interpolation to match the smallest dimension (0.24 mm). Skull stripping was deemed unnecessary, as the segmentation performance was solely evaluated within the LSA ROIs, which were distant from the skull. Additionally, experiments were conducted with and without applying N4 bias field correction from Advanced Normalization Tools (ANTs) 2.1.0 [42] to assess its impact on the segmentation performance for both DS6 and MSFDF.

##### Experiments

The 15 selected subjects were divided into three groups: a training set (N=6), a validation set (N=3), and a test set (N=6). Due to the visible variation in image quality among subjects, and to prevent the deep learning model from learning incorrect representations, we prioritised those with the highest TOF image quality for the training set. The image quality was visually rated by one rater (R.L.) based on the LSA-to-background signal contrast.

For DS6, we initially evaluated the pre-trained DS6 model as-is on our data to understand its performance without any fine-tuning. Afterwards, we fine-tuned the DS6 model on our training data, and the model state with the lowest validation loss across all training epochs was saved. We added dropout layers to the model structure to mitigate overfitting, and we conduct multiple training experiments with different dropout probabilities. The final model was selected as the one achieving the highest Dice similarity coefficient (DSC) on the validation set.

For MSFDF, separate training and validation sets were unnecessary, so they were combined into a single tuning set (N=9). We then performed a grid search through a range of pipeline parameters (see Supplementary Materials Table 4), and the optimal set was chosen as the one achieving the highest mean DSC on the tuning set.

Finally, both trained models were evaluated on the same test set. To enhance the models’ segmentation performance near the boundaries of the LSA ROIs, the segmentation was performed on the whole-brain TOF volume and then cropped to the LSA ROIs.

We assessed and compared the models’ segmentation performance within the ROIs using both quantitative and qualitative methods. Quantitatively, the segmentation was evaluated against the ground truth on DSC, balanced average Hausdorff distance (bAHD) [43], sensitivity, and positive predictive value (PPV). Both DSC and bAHD reflect the overall segmentation accuracy. Qualitatively, we visualised the segmentation in each LSA ROI on a sagittal MIP view and compared them against the ground truth using colour coding to highlight false positives and false negatives.

#### 2.2.2. Quantification Stage

This stage aims to extract the centrelines of the LSAs based on the segmentation, and to compute the final metrics of interest. It is a semi-automatic process involving both manual steps with user input and correction through the 3D Slicer software, and automatic steps performed using Python scripts. We selected 3D Slicer due to its extensive functionalities, excellent visualisation, and Python programmability.

Steps in this stage are outlined in Figure 1, and we provide a detailed user manual for this at our code repository. Briefly, the following steps are performed for each hemisphere:

1. **Define LSA ROI** (manual): the user defines an ROI box in 3D Slicer covering all LSAs and their origins.
2. **Obtain segmentation inside ROI** (automatic): the previously defined ROI is used to crop the DS6 segmentation, which can be further cleaned automatically by removing islands far from the largest one—usually the main cluster of LSAs. Additionally, different islands of this segmentation mask are colour-coded to assist manual correction.
3. **Manual correction and keep LSAs** (semi-manual): the user inspects the segmentation of LSAs and makes corrections as needed. Afterwards, only the largest island consisting of the LSAs and their originating MCA is retained, and the user cuts off the MCA.
4. **Postprocessing and detect endpoints** (automatic): the mask for each disjoint cluster of LSAs (originating from different stems) is relabelled, and its endpoints are automatically detected.
5. **Centreline extraction and construct LSA trees** (automatic): this step automatically extracts the centrelines of the LSAs, decomposes the branches, and constructs a tree of branches from each origin. It utilises the “Extract Centreline” module in the SlicerVMTK^2^ extension of Slicer.
6. **Compute LSA metrics** (automatic): based on the extracted centreline trees, we compute the final output metrics of the LSAs, which are described below.

##### Output LSA Metrics

We observed in the current literature reporting LSA morphology that, although nearly all papers reported metrics such as the length and tortuosity of LSA branches, there are inconsistent criteria for what counts as a branch [2, 13, 14], especially when there are multiple levels of bifurcation. Additionally, some studies only reported these metrics for the longest LSA observed on a 2D coronal MIP [10, 11]. Therefore, we propose to report morphological metrics for both the LSA *segments* and *branches*, where a *segment* refers to any LSA segment connecting two closest origin/terminal endpoints or bifurcation points, and a *branch* refers to the full path connecting an origin to a terminal endpoint – in an example diagram shown in Figure 3, there are 6 *segments* (A-H, H-C, H-G, G-D, G-E, B-F) and 4 *branches* (A-H-C, A-H-G-D, A-H-G-E, B-F). Besides, we defined a *stem* as the LSA segment connected to its origin on MCA/ACA.

Using the extracted LSA centrelines, our pipeline computes the following metrics:

- Number of stems
- Number of segments
- Number of branches
- Length of all segments and branches
- Tortuosity of all segments and branches – tortuosity is defined as the ratio of curved length to the straight distance connecting the two endpoints.
- Average curvature along all segments/branches – curvature is computed at each control point of the centreline and averaged. The average curvature of a segment/branch reflects how curved the segment/branch is overall, complementing the tortuosity metric.
- Maximum curvature along all segments/branches – this can reflect the presence of a sharp turn in the vessel.

**Figure 3.**
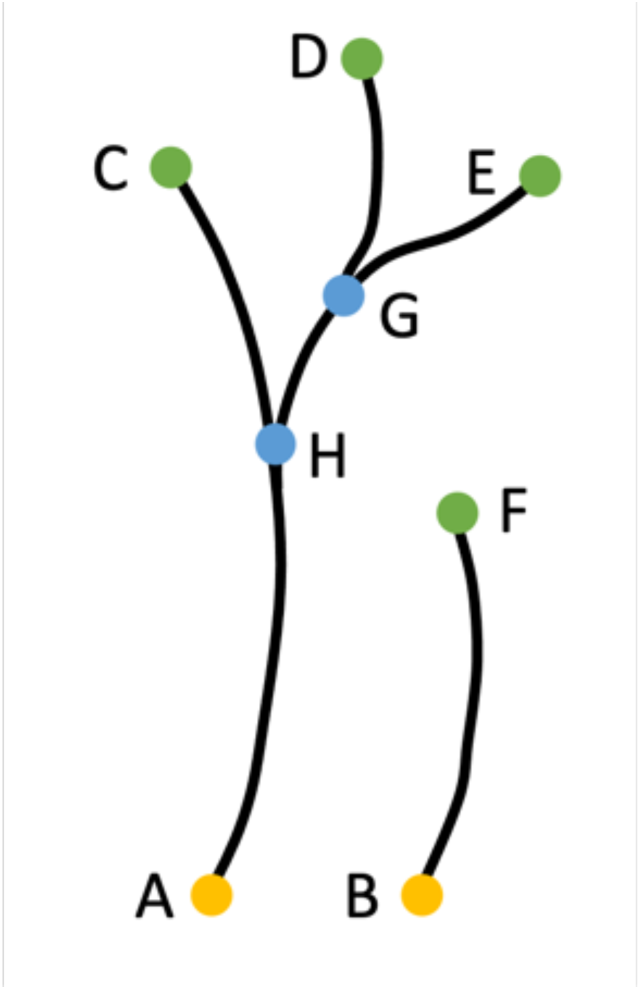
A simple diagram for the illustration of LSA segments and branches. A-B are origin endpoints, C-F are terminal endpoints, and H-G are bifurcation points.

We chose not to estimate the LSA diameter, as it was often only 1-2 voxels and is therefore highly susceptible to partial volume effects. The quantification stage was run by two trained raters (R.L. and Y.J.) on the 15 selected subjects to obtain the final metrics of their LSA morphology.

### 2.3. Comparison with 2D Analysis on Coronal MIP

As introduced previously, tracing LSAs on coronal MIP images is currently the dominant approach for analysing LSA morphology. Therefore, we sought to also conduct this analysis on the same 15 subjects and to compare the results with those obtained from our proposed pipeline.

Specifically, the MIP analysis was performed by two trained raters (S.N. and D.J.T.) using a medical image viewer software, Weasis^®^ (https://weasis.org/en/index.html). For each side of the brain, the TOF volume was reconstructed by MIP in the coronal plane with a slab thickness of 15 mm to capture the visually longest LSA branch. This selected branch was manually traced on MIP to compute its curved length and tortuosity (see Supplementary Materials Figure 1 for an example). Additionally, the numbers of LSA stems and branches in each hemisphere were counted based on both coronal and sagittal MIP images.

These extracted metrics were compared with those derived from our proposed pipeline using correlation analyses, performed on a hemisphere basis since the LSAs in each hemisphere were analysed independently in both methods. To obtain the 3D morphology of the branch selected in the MIP analysis, we located the corresponding branch by matching the terminal segment. We used Pearson’s correlation test to assess the correlation between the length and tortuosity of the selected branch measured in 2D and 3D, while Spearman’s rank correlation test was applied for the number of LSA stems and branches.

## 3. Results

### 3.1. Results for Vessel Segmentation

It was found that performing bias field correction on TOF did not impact the dice scores of the segmentation by DS6 when tested before fine-tuning, but it did improve the performance of MSFDF (Supplementary Materials Table 5). Thus, bias field correction was only kept for the MSFDF pipeline.

Results from the quantitative evaluation of the segmentation performance of the original and fine-tuned DS6, as well as of the tuned MSFDF pipeline, are shown in Table 2. We observed that fine-tuning DS6 improved its performance on all datasets and on all metrics, with the largest percentage improvement in bAHD (15-30%). The performance of the fine-tuned model on test set is comparable to that on the training and validation sets. Comparing the results for fine-tuned DS6 and MSFDF on the test set, we also observed DS6 achieving better mean DSC ↑ (0.814±0.029 vs 0.769±0.053), better bAHD **↓** (0.421±0.142 vs 0.576±0.229) and better sensitivity ↑ (0.863±0.045 vs 0.699±0.083). Nonetheless, MSFDF scored a higher mean PPV (0.865±0.036) than DS6 (0.772±0.029). Hyperparameters of the fine-tuned DS6 and MSFDF are reported in Supplementary Materials Table 4 and 6.

Qualitatively, a comparison of the segmentation by fine-tuned DS6 and MSFDF on the test subjects is illustrated in Figure 4. Red colour highlights the false-negative predictions, or the “vessels missed” in the segmentation mask, whereas blue highlights the false-positive predictions, or the “over-segmentation”. We observed that, compared with DS6, MSFDF tended to miss more branches or the terminal parts of the branches with weak signal, such as in ID 1, 4 and 6. This agrees with the lower sensitivity of MSFDF. However, in regions with extensive background noise resembling tubular structures, such as in ID 2 and 3, DS6 segmented more false positives than MSFDF.

**Figure 4.**
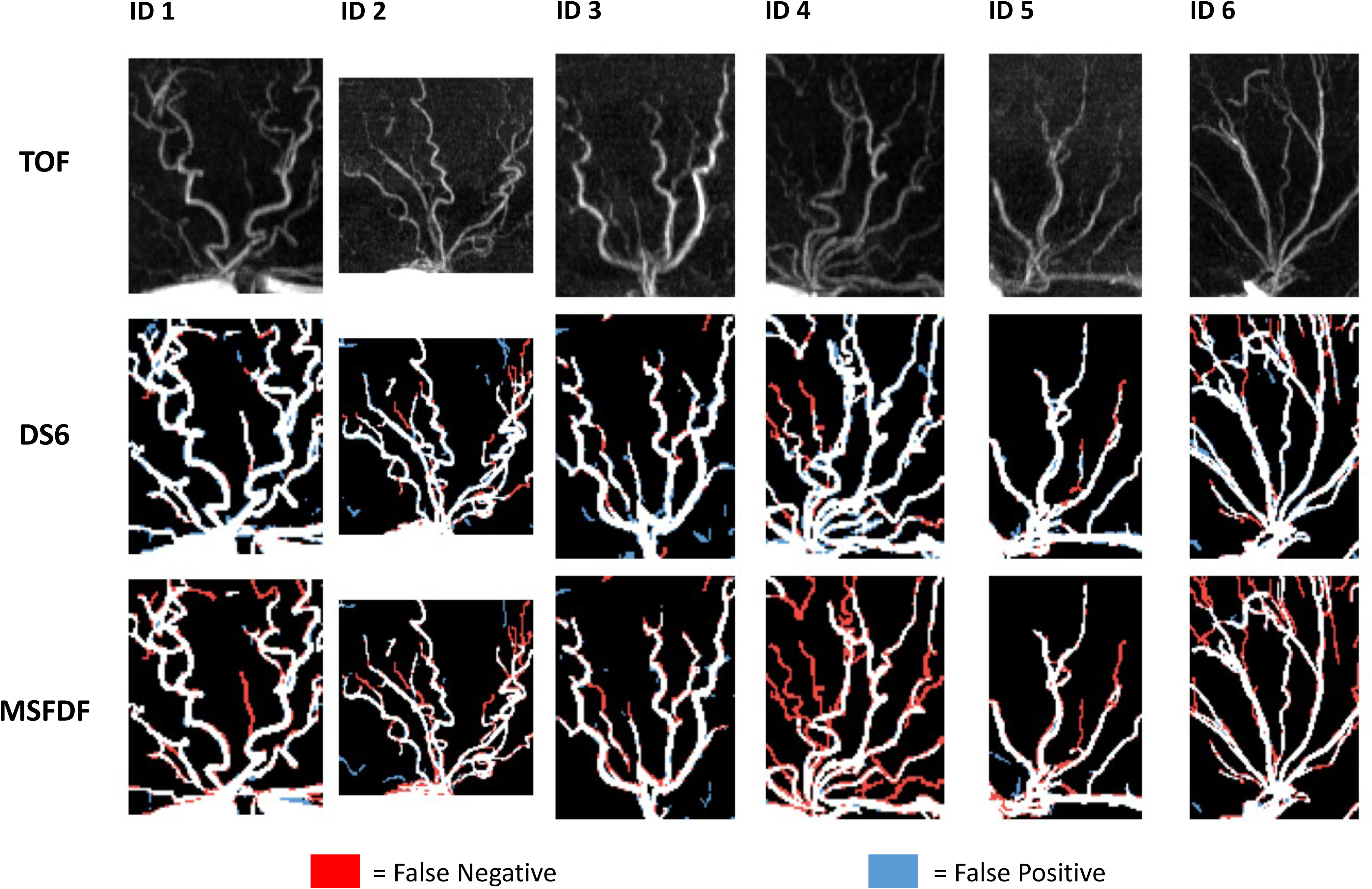
Qualitative evaluation of the segmentation by fine-tuned DS6 and MSFDF on the test subjects. Displayed are the sagittal MIP images of the LSA ROIs, showing one ROI per subject.

### 3.2. Results for LSA quantification

Figure 5 shows a few examples of the final extracted LSA centrelines, with different LSA segments colour coded. Various metrics were computed based on these extracted skeletons for the entire group of 15 subjects, and a summary of the results is reported in Table 3. On average, each subject had 5.9 ± 1.6 LSA stems, 28.7 ± 9.9 branches, and 51.3 ± 19.0 segments. The large standard deviations in the numbers of branches/segments indicated great inter-subject variations. The average length of an LSA branch was 42.5 ± 5.7 mm, and the average tortuosity was 1.86 ± 0.15.

**Figure 5.**
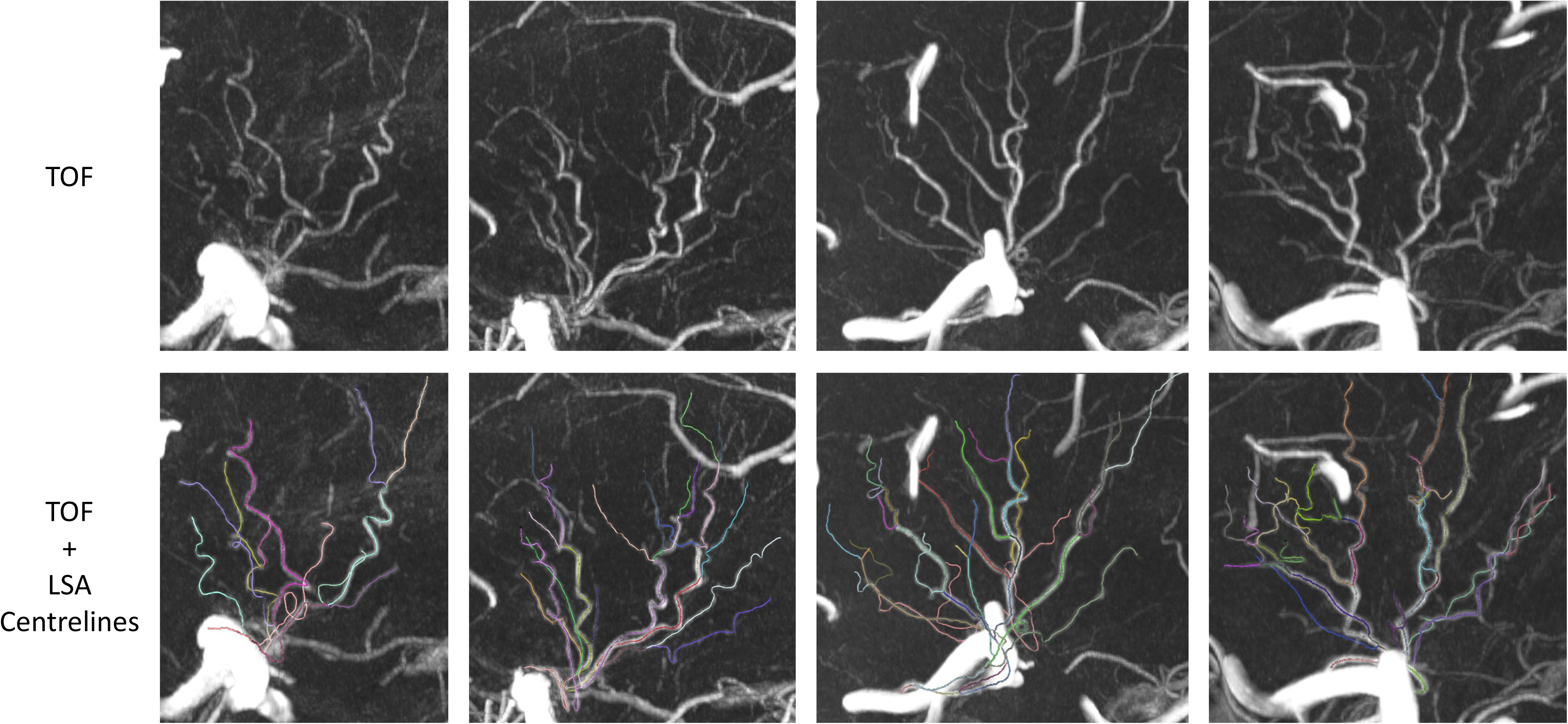
Extracted LSA centrelines and branches from our pipeline for example subjects. Different LSA segments are colour coded.

### 3.3. Correlation between LSA Morphology Evaluated in 2D and 3D

Figure 6 illustrates the correlation between the number of stems, branches, selected branch length, and tortuosity measured by the 2D MIP visual analysis and by the 3D analysis using our proposed pipeline. We also highlighted several outliers in the plots, and to help understand their values, we provide supplementary figures taken from their analyses at Supplementary Materials Figure 4.

**Figure 6.**
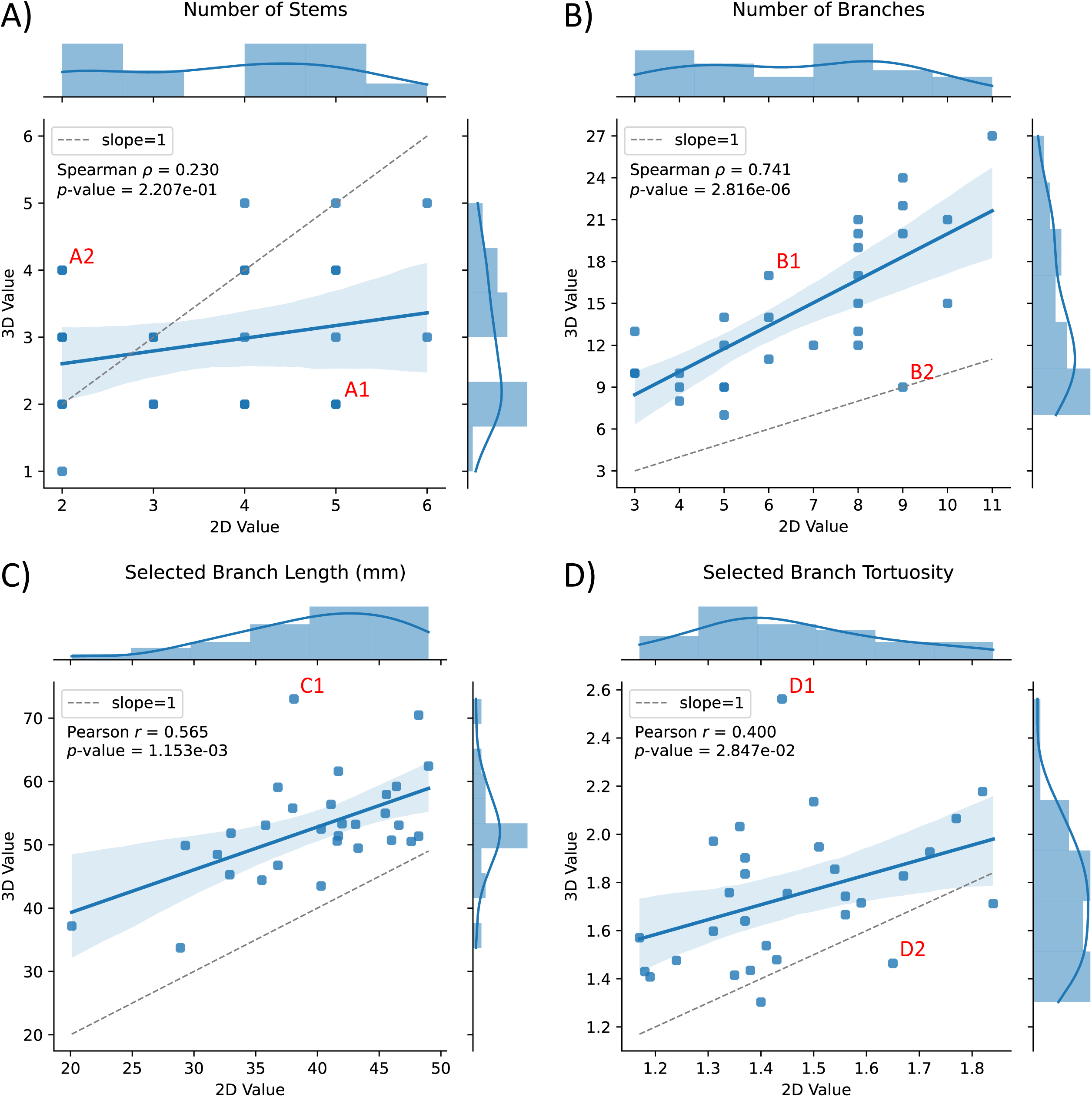
Correlation between the LSA metrics as measured by the 2D MIP analysis (2D Value) and our proposed pipeline (3D Value). Each point corresponds to one hemisphere of one subject (N=30). Some outliers are marked in red for discussion, and figures taken from their 2D and 3D analyses are provided at Supplementary Materials Figure 4.

Overall, the correlation between the 2D and 3D analysis results for the number of stems was the weakest (*ρ*=0.230, *p*=2.207e-01), and 2D analysis tended to count more stems than 3D. We found that this was because the LSA stems often locate posterior to the MCA and cannot be well visualised in either the coronal or sagittal MIPs used in the 2D analysis. For example, for outlier A1 (Supplementary Materials Figure 4-A1), there seemed to be multiple “stems” of LSAs in the coronal MIP, whereas the 3D view revealed that those “stems” were secondary segments originating from the same stem behind the MCA. On the other hand, in some cases like A2, 3D analysis identified more LSA stems originating from locations such as the ACA, which cannot be well visualised in the 2D approach (Supplementary Materials Figure 4-A2).

Unlike for the number of stems, the number of branches as counted with the two approaches showed good correlation (*ρ*=0.741, *p*=2.816e-06), though the counts from the 3D analysis were usually much higher than those from the 2D analysis due to its better visualisation of the LSAs (Supplementary Materials Figure 4-B1). In scenarios where the LSA branches are few and well separated in MIP, the two analyses arrived at the same counts (B2).

Finally, for the length and tortuosity of selected branches, their 2D values and 3D values showed only moderate correlation (length: *r*=0.565, *p*=1.153e-03; tortuosity: *r*=0.400, *p*=2.847e-02). All lengths in 3D were larger than those measured on 2D MIP as expected. The 3D tortuosity of a branch was mostly larger than that measured in 2D as well. Examining the outliers, we noticed that the 2D analysis occasionally connected disjoint LSA segments as one branch by mistake (C1), or it missed some segments near the MCA (D1). Even when an LSA branch is clearly visualized on the MIP image, it may appear far more tortuous on the coronal MIP than it actually is in 3D (D2), resulting in a significant mismatch between its 2D and 3D tortuosity.

## 4. Discussion

This paper presents a deep learning-based pipeline for analysing the 3D morphology of the LSAs in CSVD patients from contrast enhanced 7T-TOF-MRA images. It consists of an automatic vessel segmentation stage using the DS6 deep learning model, and a semi-automatic LSA quantification stage utilising custom Python code and 3D Slicer.

For the segmentation stage, we compared a state-of-the-art deep learning model, DS6, with the MSFDF pipeline based on vessel enhancement filtering, and we demonstrated the possibility to finetune DS6 with dropout on a small amount of data without overfitting. The finetuned DS6 showed a superior and more stable test performance than MSFDF in segmenting the LSAs as assessed by the quantitative metrics, and visual inspection confirmed that DS6 was more sensitive in detecting LSA branches with weak signals. The advantages we observed in this patient cohort for the deep learning model, DS6, compared to MSFDF were consistent with previous results in healthy volunteers in [30].

Moreover, unlike MSFDF, DS6 is robust to the bias field in the original TOF volume, and it does not require additional postprocessing which involves more manual tunning of the hyperparameters. Therefore, DS6 is the recommended segmentation model in our pipeline.

In clinical studies examining the morphology of LSAs, tracing LSA branches on coronal MIPs has been the dominant method due to its simplicity [2, 9, 10, 14–17], and it relies on the assumption that the 2D projection reflects the true 3D structure of the LSAs on average. However, the correlation between the results obtained through 2D MIP analysis and those from our 3D analysis offers new insights into the reliability of the 2D approach.

Specifically, we showed that using 2D MIP images is insufficient for counting the actual numbers of LSA stems and branches due to limitations in the visualisation angles. Nonetheless, the branch counts in 2D correlate relatively well with those counted in 3D. The 2D and 3D measurements of the length and tortuosity of the longest branch selected in the 2D analysis only exhibited moderate correlation, with the 3D values often spanning over a larger range. This indicates that they may correlate differently with other clinical features of the patients, too, and that the 3D measurements might be more sensitive in detecting changes. Furthermore, analysis of the outliers showed how tracing the LSA branch on 2D may mistakenly connect disjoint segments due to the visualisation angle, thus making the results less reliable. These observations highlight the need to assess the morphology of LSAs in 3D.

The strengths of our study include being, to the best of our knowledge, the first study to apply deep learning techniques to the segmentation of LSAs. Unlike many studies that performed vessel segmentation without assessing the accuracy quantitatively [11, 13, 21, 22], we also performed a rigorous evaluation of the segmentation against manually labelled ground truth on a voxelwise basis. Our manually expert labelled ground truth is available on reasonable request for future studies. Moreover, we assessed the correlation between the LSA morphology measured on MIP and that measured in 3D, and we have not seen similar analyses in the existing literature.

However, our study has some limitations. We developed and evaluated our pipeline using only 15 subjects, constrained by the time-intensive process of manually creating ground truth masks for each subject (∼10hr/subject). Our pipeline is also not fully automatic and requires some manual input, such as filling the segmentation of LSA terminals with very weak signals, particularly when image quality is marginal. Nonetheless, our approach reduces the burden of manual input to a level where 3D segmentation and vessel morphological analysis is now feasible. Moreover, in some TOF volumes, we observed significant pulsation-related artifacts near the MCAs, which complicated segmentation of the LSA origins and may have introduced inaccuracies in the extracted morphological metrics. Thus, future work may consider only examining the LSAs inside the basal ganglia area, or the on-scanner sequences could be updated to reacquire these key slices during the patient scan should they be corrupted by pulsatile artefacts to improve the robustness of LSA segmentation.

## 5. Conclusions

We present a semi-automatic pipeline for evaluating the 3D morphology of LSAs in patients with CSVD based on contrast enhanced 7T-TOF-MRA data. We applied deep learning to LSA segmentation for the first time and achieved a mean DSC of 0.814 in testing, outperforming the classical MSFDF pipeline by 5.5%. The final outputs from our pipeline include the numbers of LSA stems, segments and branches, along with measurements of LSA length, tortuosity, and curvature. Compared with 2D MIP analysis, our pipeline provides a more truthful characterisation of the 3D morphology of LSAs, which may be more sensitive to CSVD progression. Our open-source pipeline is designed to support clinical studies aimed at characterising the 3D morphology of LSAs in CSVD, paving the way for a deeper understanding of its pathophysiology.

## Data and Code Availability

Our pipeline was developed with 3D Slicer version 5.7.0-2024-01-28, and Python 3.10.13. The finetuned DS6 model is available at https://huggingface.co/soumickmj/SMILEUHURA_DS6_CamSVD_UNetMSS3D_wDeform (retrieved on 20/09/2024). Code for our pipeline and detailed instructions are available at https://github.com/RuiLiGitLove/LSA_3D_Morph (retrieved on 20/09/2024).

## Acknowledgements

This work was supported by a British Heart Foundation project grant [PG/19/74/34670]. HSM’s research receives Infrastructural support from the Cambridge British Heart Foundation Centre of Research Excellence [RE/24/130011]. HSM and CTR are supported by the Cambridge University Hospitals NIHR Biomedical Research Centre [NIHR203312]. The views expressed in this publication are those of the authors and not necessarily those of the NIHR, NHS, or UK Department of Health and Social Care. CTR receives research support from Siemens Healthcare for a different project. The 7T MRI was supported by an MRC Clinical Research Infrastructure Award [MR/M008983/1]. RL was supported by a PhD studentship awarded by Trinity College, University of Cambridge, UK. These funding sources were not involved in the study design, data collection, analysis, interpretation of data, writing, or manuscript submission for this study. For the purpose of open access, the author has applied a CC BY public copyright licence to any Author Accepted Manuscript version arising from this submission.

## Author Contributions

**Rui Li:** conceptualisation, methodology, software, formal analysis, investigation, data curation, writing-original draft, visualisation.

**Soumick Chatterjee:** methodology, writing – review & editing, supervision

**Yeerfan Jiaerken:** methodology, validation, data curation, investigation

**Chethan Radhakrishna:** software

**Philip Benjamin:** methodology, data curation

**Stefania Nannoni:** investigation

**Daniel J. Tozer:** methodology, investigation, formal analysis, data curation, writing – review and editing.

**Hugh S. Markus:** conceptualisation, methodology, funding acquisition, supervision, writing – review & editing.

**Christopher T. Rodgers:** conceptualisation, methodology, software, investigation, resources, writing – review and editing, supervision

## Footnotes

1 Retrieved on 20/09/2024. Commit ID=1df8bc8.

2 https://github.com/vmtk/SlicerExtension-VMTK. Retrieved on 20/09/2024.

