## Supplementary Materials for "A Deep Learning Pipeline for Analysis of the 3D Morphology of the Cerebral Small Perforating Arteries from Time-of-Flight 7 Tesla MRI"

**Figures**

**Figure 1.** The maximum intensity projection image in the coronal plane of an example subject from our cohort, with manual tracing of an LSA branch.

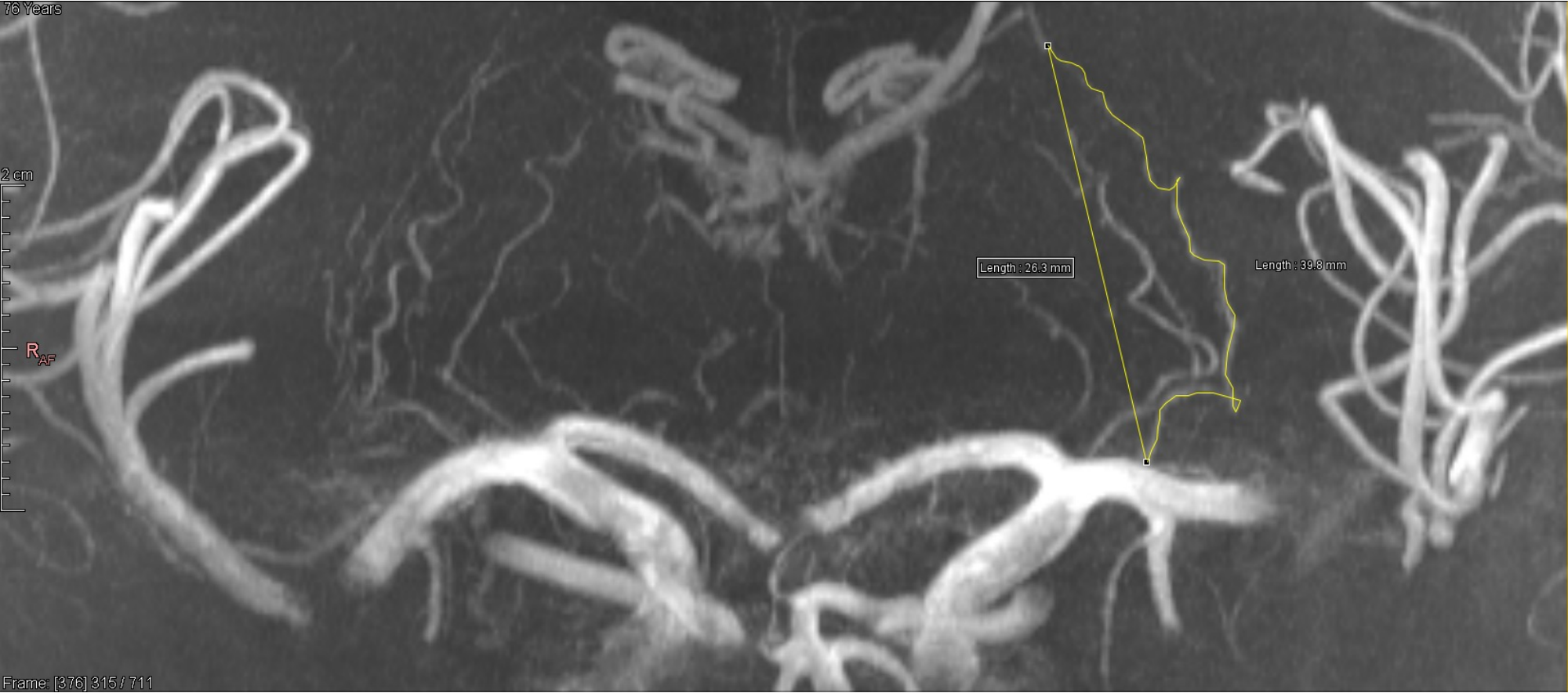

**Figure 2.** Diagram of the MSFDF pipeline.

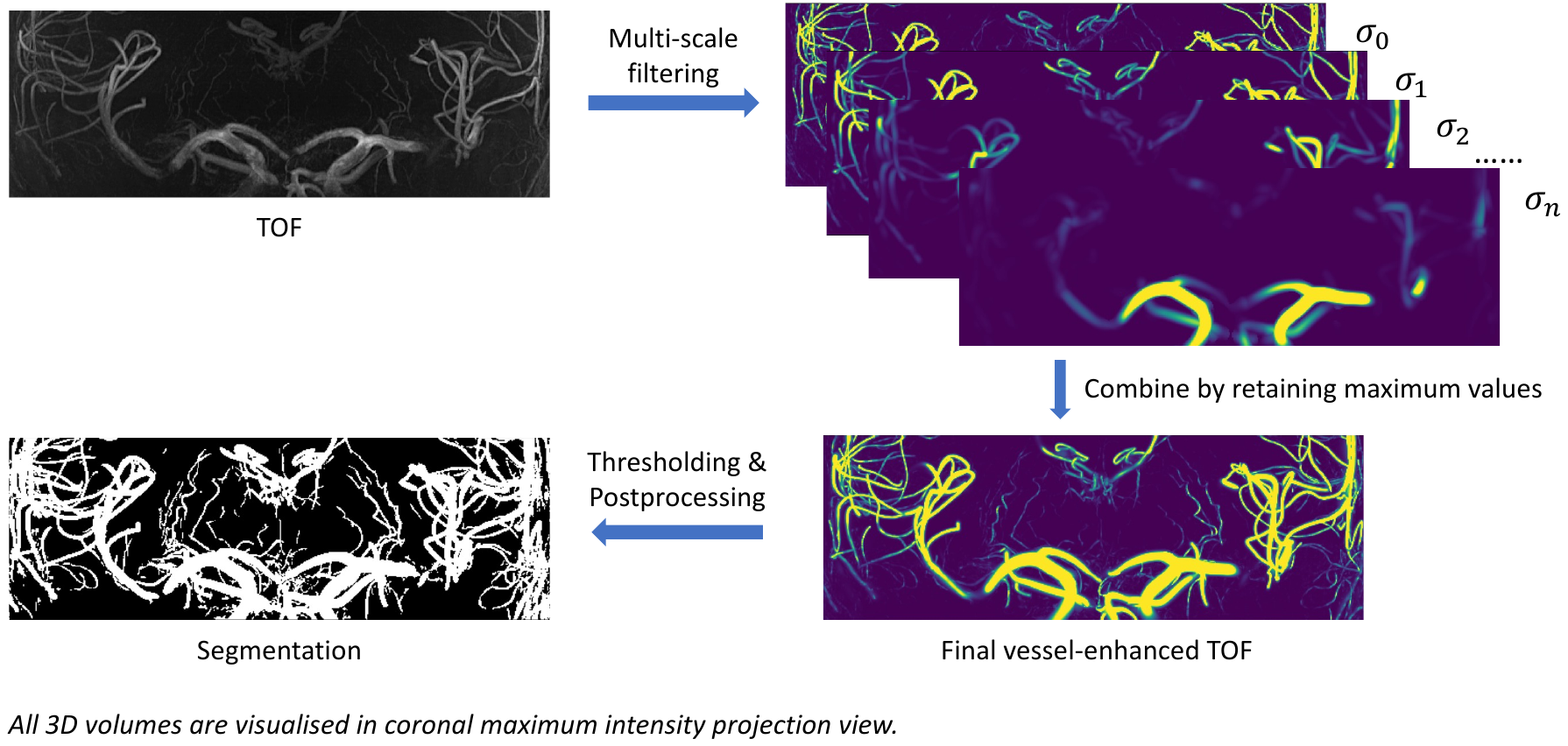

**Figure 3.** Segmentation for an example subject produced by the original MSFDF pipeline with Otsu thresholding.

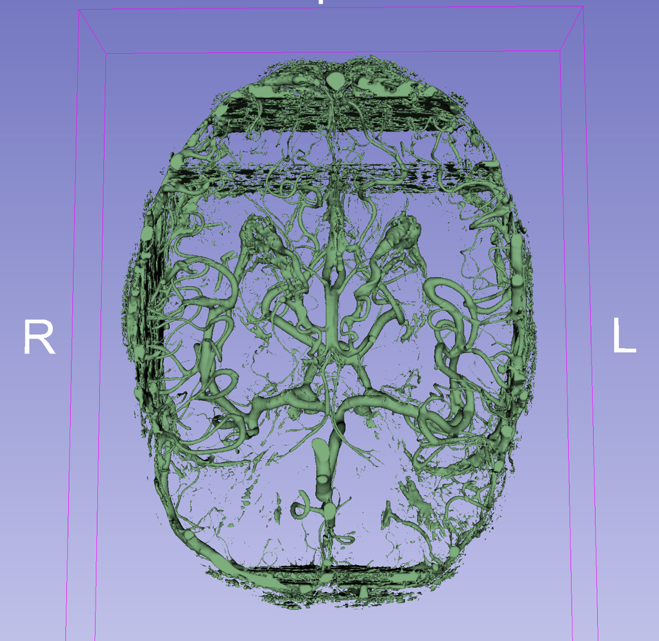

**Figure 4.** Figures taken from the 3D and 2D analyses for outliers in the correlation plots.

**A1** – Number of stems; 3D value=2, 2D value=5.

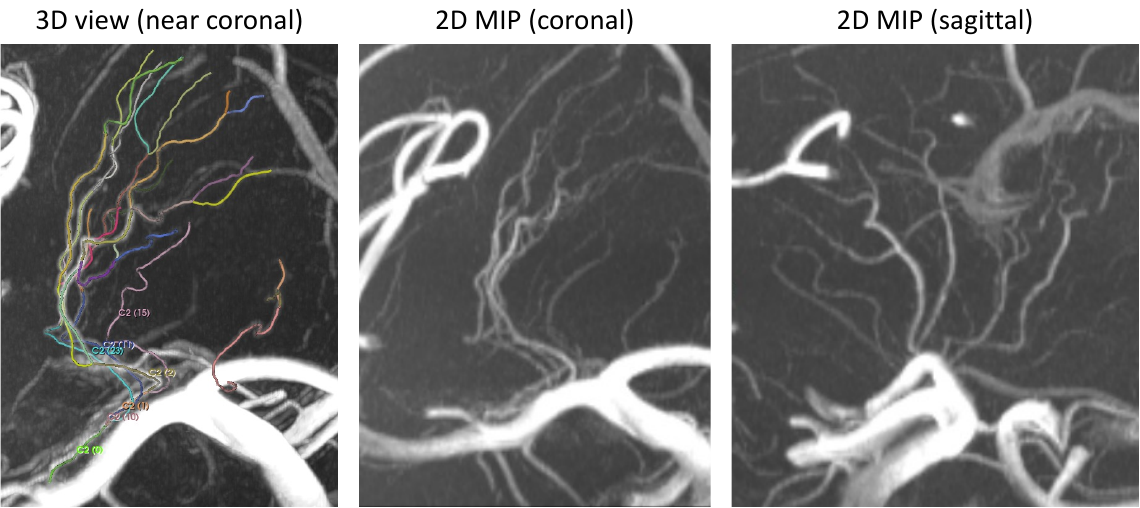

**A2** – metric=number of stems; 3D value=4, 2D value=2.

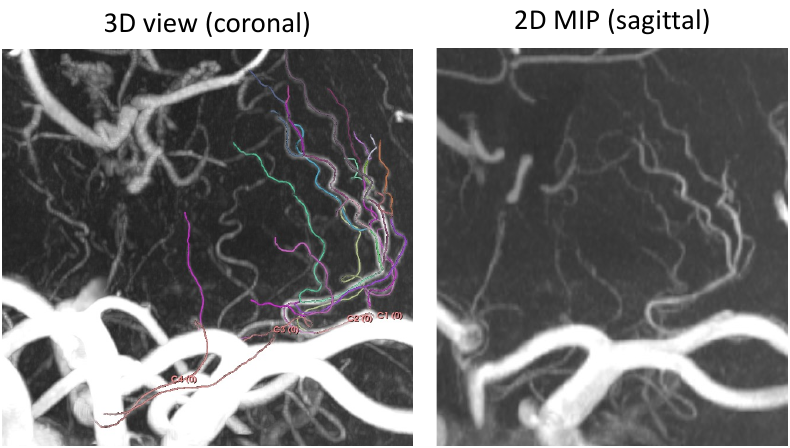

**B1** – metric=number of branches; 3D value=17, 2D value=6.

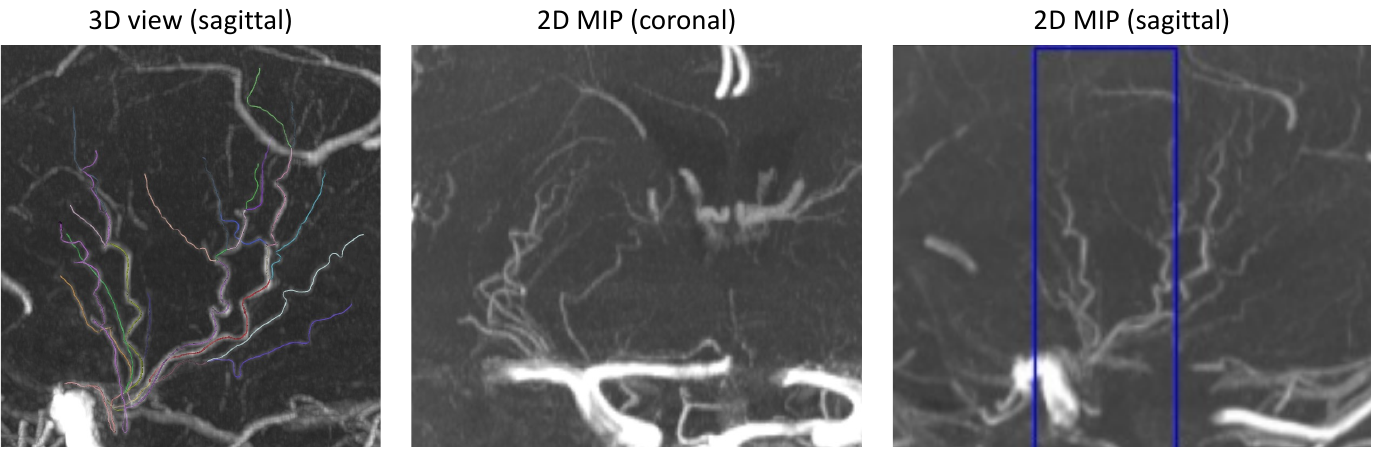

**B2** – metric=number of branches; 3D value=9, 2D value=9.

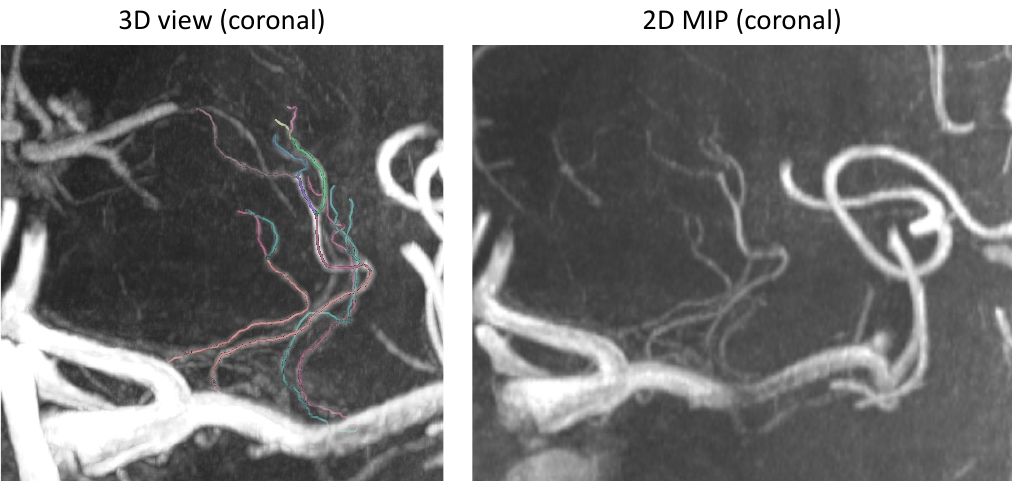

**C1** – metric=selected branch length; 3D value=73.02 mm, 2D value=38.1 mm.

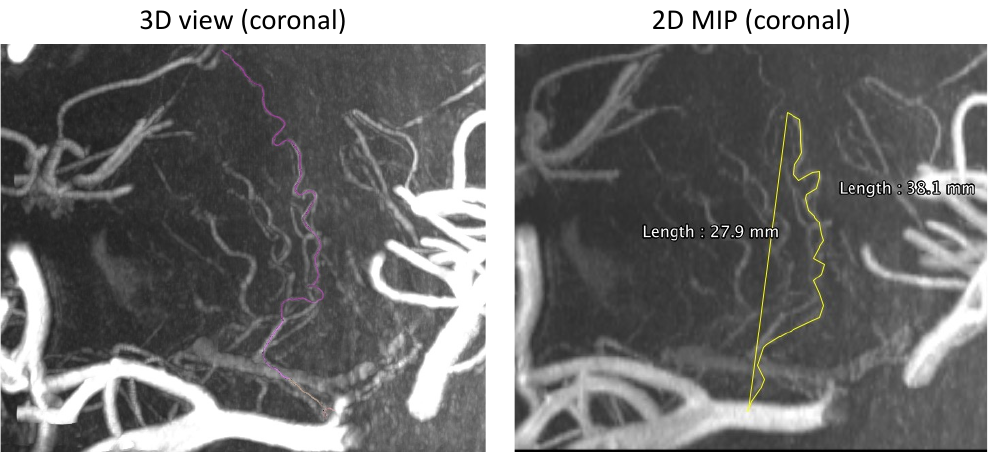

**D1** – metric=selected branch tortuosity; 3D value=2.56, 2D value=1.44.

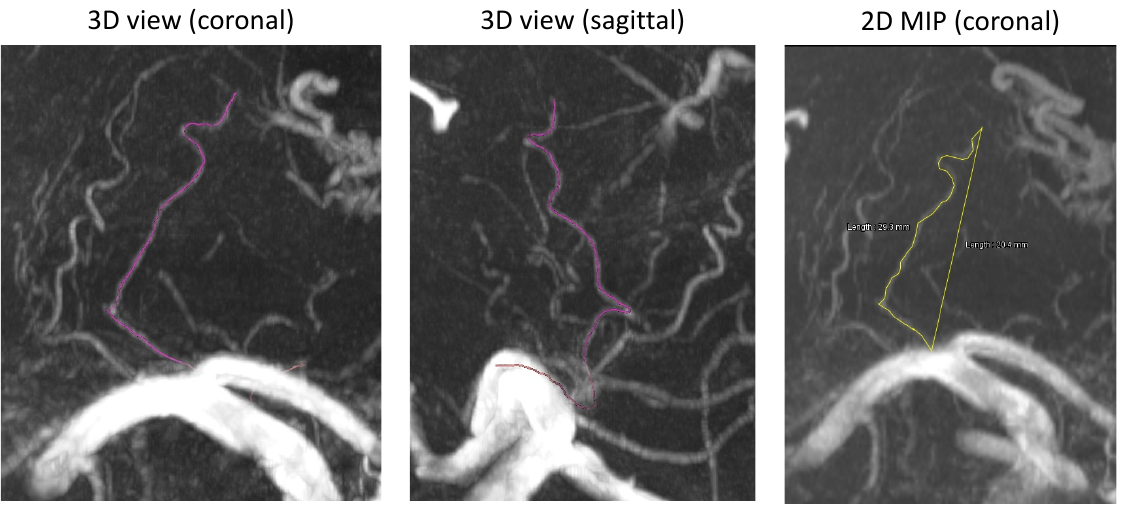

**D2** – metric= selected branch tortuosity; 3D value=1.46, 2D value=1.65.

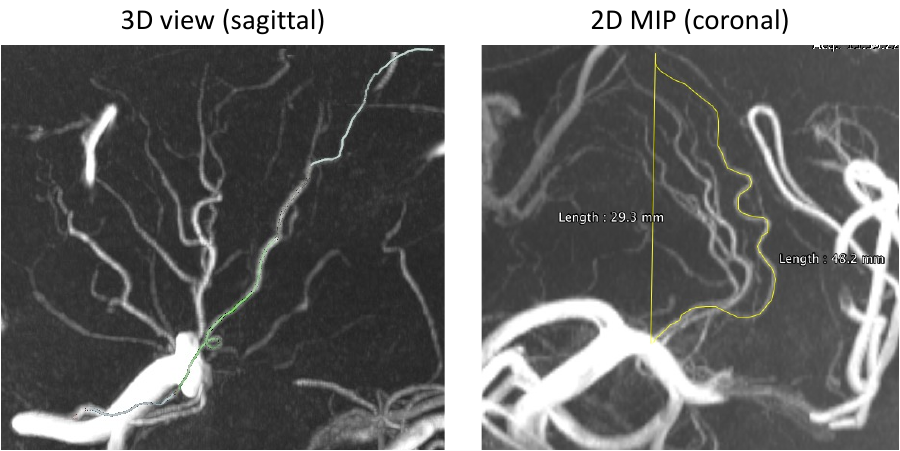

**Tables**

**Table 1.** Summary table of related work

| Authors | Year | Method Summary | Imaging Type | Subjects | 2D/3D Analysis | LSA Segmentation | Morphology Quantification | Code |
| --- | --- | --- | --- | --- | --- | --- | --- | --- |
| Li *et al.*  Wei *et al.*  Xu *et al.*  Osuafor *et al.*  Yashiro *et al.*  Xie *et al.*  Jiang *et al.* [1–7] | Multi | Manual labelling on 2D MIP | TOF,  vessel-wall imaging | Various patient groups | 2D | ✕ | ✓ | ✕ |
| Liao *et al.* [8] | 2016 | 3D tube fitting + gap filling by minimal path finding | TOF | Vascular dementia patients | 3D | ✓ (but did not evaluate accuracy) | ✓ | ✕ |
| Bernier *et al.* [9] | 2018 | Multi-Scale Frangi Diffusive Filter (MSFDF) pipeline | TOF,  SWI | Young healthy volunteers | 3D | ✕ (vasculature) | ✕ | ✓ |
| Liu *et al.* [10] | 2021 | Modified Frangi filtering with automatic seed detection and region growing | TOF | Young healthy volunteers | 3D | ✕ (vasculature) | ✕ | ✕ |
| Deshpande *et al.* [11] | 2021 | Frangi filtering | TOF, CTA | Healthy volunteers, stroke patients | 3D | ✕ (vasculature) | ✕ | Available upon request |
| Wei *et al.* [12] | 2021 | Vessel enhancement filtering | TOF | Healthy volunteers | 2D, 3D | ✓ (but did not evaluate accuracy) | ✓ | ✕ |
| Livne *et al.* [13] | 2019 | 2D U-Net segmentation | TOF | Cerebrovascular disease patients | 3D | ✕ (vasculature) | ✕ | Available upon request |
| Fan *et al.* [14] | 2019 | Hidden Markov random field + U-Net | TOF | Healthy volunteers, stroke patients | 3D | ✕ (vasculature) | ✕ | ✕ |
| Chatterjee *et al.* [15] | 2022 | “DS6” – 3D U-Net + multi-level deep supervision + elastic deformation consistency learning | TOF | Healthy volunteers | 3D | ✕ (vasculature) | ✕ | ✓ |
| Tetteh et al. [16] | 2020 | Fully convolutional network with 3D cross-hair filters | TOF, CTA | Unspecified | 3D | ✕ (vasculature) | ✓ | ✓ |

TOF = time-of-flight; SWI = susceptibility weighting imaging; CTA = computed tomography angiography.

**Table 2.** Inclusion and exclusion criteria for participant recruitment

| **Group** | **Inclusion** | **Exclusion** |
| --- | --- | --- |
| Non-acute lacunar strokes with SVD | Evidence of SVD defined as at least one of the following:   1. A lacunar stroke syndrome (e.g., pure motor stroke, pure sensory stroke, sensorimotor stroke, ataxic hemiparesis or clumsy hand dysarthria syndrome) with an anatomically corresponding lacunar infarct on CT brain scan or T1, fluid-attenuated inversion recovery (FLAIR) or diffusion weighted imaging (DWI) MRI of mild (NIHSS score ≤ 8) or moderate (NIHSS score 9-15) severity [17]; 2. Vascular cognitive impairment associated with MRI features of SVD; 3. Gait apraxia associated with MRI features of SVD; 4. A genetic diagnosis of a monogenic form of SVD e.g., CADASIL. | 1. Unable/unwilling to consent; 2. Age<18; 3. Stroke aetiology due to cardio-embolism (as defined according to the TOAST criteria [18]) or large vessel disease (>50% stenosis in extra- or intra-cranial cerebral arteries on NASCET criteria [19]); 4. Lacunar infarcts > 1.5cm – as many of these infarcts are caused by embolism; 5. Severe stroke (NIHSS score ≥ 16) [17]; 6. Evidence of cortical infarct of any size; 7. Other major neurological diseases; 8. Severe systemic diseases such as heart failure, liver failure and kidney failure or any illness in the judgement of the investigator that could affect participation in the study; 9. MRI contraindications e.g., metal objects in or on the body, claustrophobia, pregnancy, known allergy to gadolinium containing contrast agent, impaired renal function with estimated glomerular filtration rate (eGFR) <59ml/min/1.73m^2^. |
| Healthy volunteer controls | 1. Stroke free subjects of similar age to patient groups; 2. No evidence of other major neurological diseases. | 1. Unable/unwilling to consent; 2. Age<18; 3. History of stroke or other major neurological illness; 4. MRI contraindications e.g., metal objects in or on the body, claustrophobia, pregnancy, known allergy to gadolinium containing contrast agent, impaired renal function with estimated glomerular filtration rate (eGFR) <59ml/min/1.73m^2^. |
| Non-acute stroke controls without SVD | Evidence of non SVD related stroke defined as:  A partial or total anterior circulation stroke syndrome (e.g., Hemiparesis with or without hemisensory loss, hemianopia, dysphasia or neglect) suspected to be due to large artery atherosclerosis or cardio-embolism of mild (NIHSS score ≤8) or moderate (NIHSS score 9-15) severity. | 1. Unable/unwilling to consent; 2. Age<18; 3. Stroke aetiology due to small vessel disease/small artery occlusion; 4. An extracranial or intracranial internal carotid artery occlusion or a proximal (M1) middle cerebral artery occlusion which would be more appropriately treated with combined intravenous intra-arterial therapy; 5. Severe stroke (NIHSS score ≥16) [17]; 6. Intracranial haemorrhage identified on initial CT; 7. Other major neurological diseases; 8. Severe systemic diseases such as heart failure, liver failure and kidney failure or any illness in the judgement of the investigator that could affect participation in the study; 9. MRI contraindications e.g., metal objects in or on the body, claustrophobia, pregnancy, known allergy to gadolinium containing contrast agent, impaired renal function with estimated glomerular filtration rate (eGFR) <59ml/min/1.73m^2^. |

**Table 3.** MSFDF segmentation performance on the tuning dataset with and without post-processing. Reported are the mean and standard deviation of the dice scores.

| Model | Dataset | No post-processing | With postprocessing |
| --- | --- | --- | --- |
| MSFDF | Tuning | 0.763 (0.075) | 0.777 (0.065) |

**Table 4.** Search range and optimal parameters for the MSFDF model.

| Parameter | Search Range | Optimal Value |
| --- | --- | --- |
| Sigma minimum | [1] | 1 |
| Sigma maximum | [7,8] | 7 |
| Gamma | [300] | 300 |
| Local threshold offset | [-0.01, -0.02] | -0.01 |
| Local threshold block size | [25] | 25 |
| Minimum cluster size | [100, 120, 140, 160, 180, 200, 220] | 180 |

**Table 5.** Segmentation performance of the original DS6 model and MSFDF pipeline on different datasets with and without performing bias field correction on the input TOF image. Presented are mean and SD of dice similarity coefficients on each dataset.

| Model | Dataset | No Bias Field Correction | With Bias Field Correction |
| --- | --- | --- | --- |
| DS6 | Training | 0.811 (0.057) | 0.811 (0.056) |
|  | Validation | 0.780 (0.053) | 0.780 (0.053) |
| MSFDF | Tuning | 0.767 (0.078) | 0.777 (0.065) |

**Table 6.** Training hyperparameters for the fine-tuned DS6 model.

| Hyperparameter | Value |
| --- | --- |
| Learning rate | 0.000001 |
| Total epochs | 50 |
| Dropout rate | 0.25 |
| Samples per epoch | 8000 |
| Patch size | 64 |
| Stride length | 16 |
| Stride width | 16 |
| Stride depth | 8 |
